# A software package for simple and rigorous survival machine learning analysis in biomedical research

**DOI:** 10.64898/2026.08.05.26359034

**Authors:** Alyssa F. Pybus, Junhao Qiu, Paulo Cilas Morais Lyra, Khai Dang, Isis Narvaez-Bandera, Tosin Jolaogun, Jeremy Goecks

**Author notes:** Corresponding author, (JG).

## Abstract

Survival analysis is a fundamental technique in biomedical research for modeling time-to-event data. It enables the identification of prognostic factors in disease, compares survival outcomes across treatment groups, and performs targeted treatment selection. A variety of machine learning (ML) approaches to survival analysis have emerged to complement classical statistical methods, especially for high-dimensional datasets with complex, nonlinear interactions between features. However, using survival ML methods requires addressing challenges such as censoring-unaware evaluation, overfitting, selecting performance metrics, and data leakage. To address these and other difficulties in using survival ML models, we developed the `mlsurv` software package. `mlsurv` is an open-source Python package built around three major design principles: 1) methodological rigor, including evidence-based model selection, leakage-free pipelines, and multi-metric evaluation, 2) multi-scale evaluation and interpretation, including population and subpopulation evaluation, patient-level explanations, and feature analysis, and 3) automated trust and transparency, including limitation flagging and TRIPOD+AI-aligned reporting. `mlsurv` bundles ten models spanning linear, ensemble, kernel, and deep learning families within a unified software package. To our knowledge, mlsurv is the first package to span the complete survival ML workflow from automated model recommendation through TRIPOD+AI reporting and individual patient explanation. We demonstrate `mlsurv` on the Chowell immunotherapy cohort (n=1,479). We found that overall survival (OS) was more predictable than progression-free survival (PFS) (concordance of 0.73 vs 0.67). Albumin was a top feature for both endpoints but dominated OS prediction, whereas tumor mutational burden rose to co-lead PFS prediction. Survival models matched the response-trained LORIS clinical score on PFS prediction and exceeded it on OS. `mlsurv` enables biomedical researchers to conduct rigorous, multi-model survival analysis and benchmarking using minimal code with default best practices rather than implementing custom scripts and methodological safeguards from scratch.

## Background

Survival analysis underpins prognosis, treatment selection, and trial design across biomedical research and clinical practice. Survival methods analyze the time until an event of interest (e.g., progression, death) and account for censored observations where event time is unknown beyond a subject’s last follow-up. Survival and hazard functions summarize event risk over time for each patient and are traditionally modeled with covariates via Cox proportional hazards regression (1). However, modern biomedical datasets often feature high dimensionality, complex feature interactions, and non-proportional hazards that Cox regression does not accommodate.

Researchers are increasingly adopting machine learning (ML) for survival prediction in modern complex biomedical datasets (2–7). However, executing robust ML survival analysis requires overcoming several challenges. First, ML models must be selected, tuned, and evaluated to ensure methodological rigor and avoid leakage as well as overfitting that can inflate reported performance (8). Second, comprehensive model evaluation and interpretation are required at different levels of observation, from subpopulations to individuals, and across features and their interactions. Third, best practices are needed to promote trustworthiness of the analysis, including verifying model assumptions and dataset adequacy, enabling reproducibility, and comprehensive reporting. The growing use of artificial intelligence (AI) to perform biomedical analyses further amplifies these concerns, as such tools may omit steps or introduce errors (9,10).

Existing software tools address isolated parts of this workflow, but none provide an integrated pipeline spanning model selection through patient-level explanation. Current software tools perform parametric modeling (11,12), machine learning models (12–14), deep learning (15,16), and hyperparameter tuning (14,17), but rarely encompass a full survival analysis from start to finish. This fragmentation carries real methodological risks. Data leakage remains the most common error in ML research, affecting a substantial fraction of published studies (18,19).

Model selection itself lacks clear guidance, as no single model class reliably dominates across survival datasets (20,21). For example, a recent meta-analysis of 21 cancer survival studies found extensive between-study heterogeneity in ML vs. CoxPH performance (22), suggesting task-specific model benchmarking is essential for performance optimization. Despite this evidence, ad hoc analyses routinely commit to a specific model without systematic comparison.

To bridge these gaps and enable rigorous, comprehensive ML-based survival analysis, we have developed `mlsurv`, an open-source software package for robust, end-to-end ML survival modeling. `mlsurv` is implemented in Python using three design principles: methodological rigor, multi-scale evaluation and interpretation, and automated trust and transparency (**Table 1**). `mlsurv` bundles ten survival analysis models from multivariate Cox proportional hazards (CoxPH) to deep learning models behind a unified tool with built-in methodological safeguards. Each experiment is organized as a single end-to-end pipeline with discrete analytical stages: leakage-protected preprocessing, evidence-based model recommendation, hyperparameter tuning, prospective patient risk scoring with bootstrap confidence intervals, population-, subpopulation-, and patient-level interpretation via SHAP explanations and feature-interaction analysis, and TRIPOD+AI (Transparent Reporting of a multivariable prediction model for Individual Prognosis Or Diagnosis, plus Artificial Intelligence) reporting. We validate `mlsurv` by reproducing published survival analyses across multiple models and datasets. We then demonstrate the full `mlsurv` workflow on the Chowell immunotherapy cohort (23) as an in-depth case study to demonstrate the complete workflow. In this cohort, survival-trained models match or exceed the performance of the response-trained LORIS biomarker, highlighting the differences between overall and progression-free survival prediction as well as prognostic versus predictive biomarkers.

**Table 1.** Three design principles for biomedical survival machine learning and their implementation in mlsurv.

| Design Principle | Risk Addressed | `mlsurv` Solution |
| --- | --- | --- |
| Methodological rigor | Overfitting, data leakage, and evaluation errors that inflate reported performance | Data-driven model selection guidance, leakage-free preprocessing, and censoring-aware multi-metric evaluation |
| Multi-scale evaluation and interpretation | Summary metrics obscure subpopulation disparities, feature drivers, and individual risk factors | Evaluation across population, subpopulation, and individual scales; feature importance, interaction, and local explanation analysis |
| Automated trust and transparency | Undetected statistical issues, no audit trail, and gaps in TRIPOD+AI compliance | Automated limitation flagging, structured interactive reporting, and TRIPOD+AI compliance scaffolding |

## Implementation

The sections below highlight the main features and innovations of `mlsurv.` Detailed software reference, usage examples, and implementation notes for `mlsurv` are available at https://github.com/goeckslab/mlsurv/tree/main/docs.

### Overview

`mlsurv` makes it simple to perform survival ML analyses by requiring minimal code to perform a complete analysis using best practices. The package provides a set of high-level commands for an entire survival machine learning workflow through pipeline configuration, training, evaluation, prediction, analysis, and reporting (**Fig 1**). An entire workflow can be completed with just three commands – `setup()`, `run()`, and `generate_report()` – while optional methods are available for customizable workflows and in-depth analysis. `setup()` configures the preprocessing pipeline, cross-validation strategy, and train/test split. `run()` handles the full modeling cycle from training and tuning through evaluation and feature analysis, returning structured results. Lastly, `generate_report()` assembles all results into an interactive report. **S1 Table** lists the full set of high-level functions provided by `mlsurv`.

**Fig 1.**
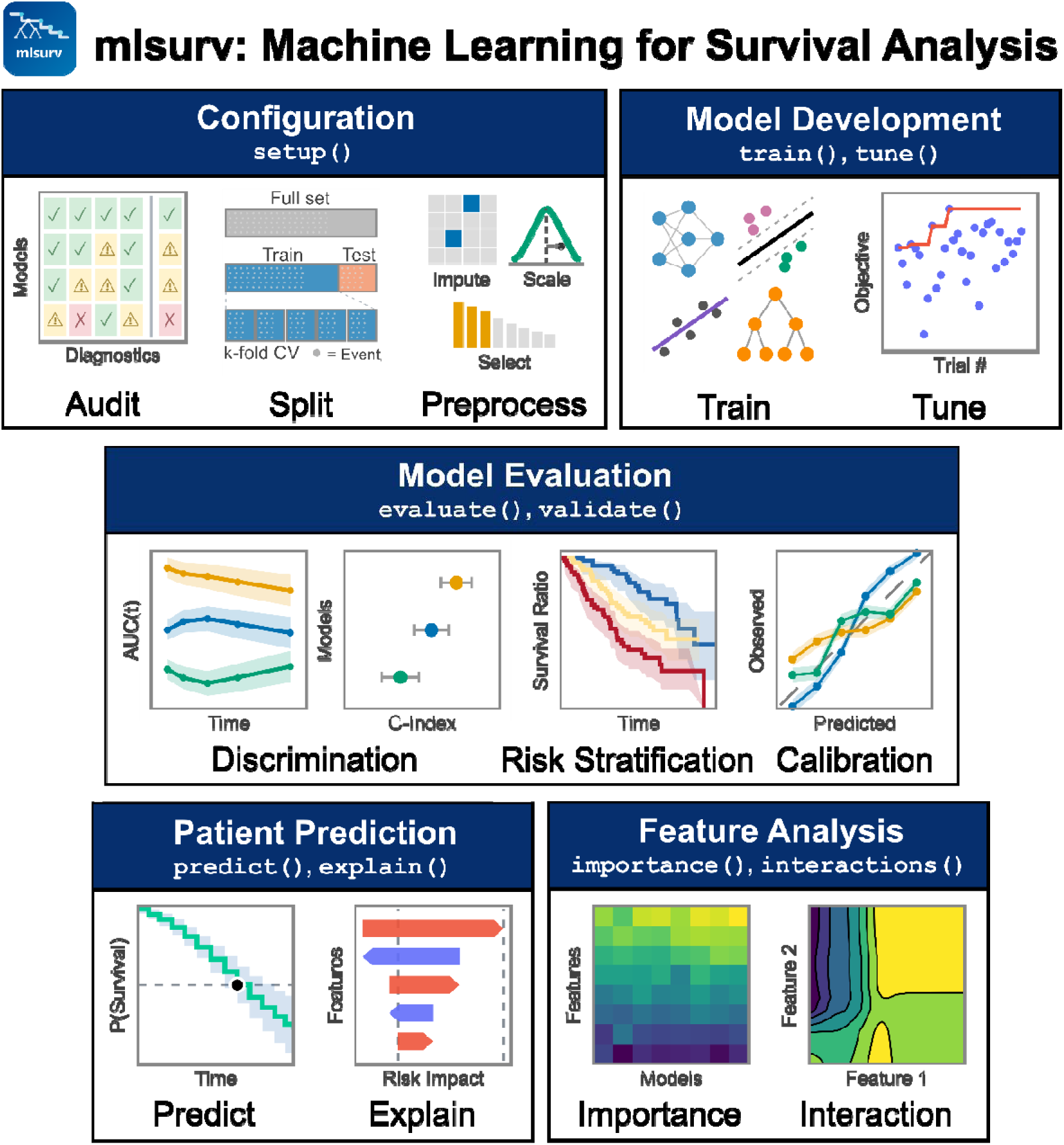
Overview of the mlsurv software package. Stages of a standard survival ML workflow are listed in each table alongside their corresponding `mlsurv` methods. During configuration, the `setup()` command audits the dataset to recommend appropriate models, splits the dataset into test/train and CV folds, and sets preprocessing steps. In model development, the `train()` command trains a variety of survival ML models and `tune()` conducts hyperparameter optimization. Model evaluation is performed by the `evaluate()` and `validate()` commands to calculate discrimination and calibration metrics as well as conduct risk stratification. Survival prediction on new patient data is performed with the `predict()` command. SHAP risk explanation by feature is computed by `explain()`. Feature analysis is performed with the `importance()` and `interactions()` commands. `mlsurv` integrates ten survival models spanning four families of analysis: linear, ensemble, kernel, and deep learning (**Table 2**). All models share a unified interface for fitting, prediction, survival function estimation, scoring, and feature importance extraction.

**Table 2.** Survival models available in mlsurv.

| Model | Abbrev. | Family | PH Assum. | Source | Reference |
| --- | --- | --- | --- | --- | --- |
| Cox Proportional Hazards | CoxPH | Linear | Yes | scikit-survival (12) | Cox (1972) (1) |
| Elastic Net Cox Regression | CoxNet | Linear (regularized) | Yes | scikit-survival (12) | Simon et al. (2011) (24) |
| Weibull Accelerated Failure Time | Weibull AFT | Linear (parametric) | Yes | lifelines (11) | Kalbfleisch & Prentice (2002) (25) |
| Random Survival Forest | RSF | Ensemble | No | scikit-survival (12) | Ishwaran et al. (2008) (26) |
| Gradient Boosted Survival Analysis | GBSA | Ensemble | Yes* | scikit-survival (12) | Hothorn et al. (2006) (27) |
| XGBoost Cox Regression | XGBoost-Cox | Ensemble | Yes | XGBoost (13) | Chen & Guestrin (2016) (13) |
| XGBoost Accelerated Failure Time | XGBoost-AFT | Ensemble | No | XGBoost (13) | Barnwal et al. (2022) (28) |
| Fast Survival Support Vector Machine | FSSVM | Kernel | No | scikit-survival (12) | Pölsterl et al. (2015) (29) |
| DeepSurv | DeepSurv | Deep Learning | Yes | pycox (15) | Katzman et al. (2018) (30) |
| DeepHit | DeepHit | Deep Learning | No | pycox (15) | Lee et al. (2018) (31) |

\*GBSA assumes proportional hazards (PH) only with the default Cox partial-likelihood loss.

### Data and Pipeline Configuration

`mlsurv` loads survival data from tabular files and validates event encodings, rejecting non-positive times. `setup()`configures the train/test split, cross-validation (CV) strategy, and preprocessing components. Default settings use a 30% event-stratified test split, five-fold CV, and Harrell’s concordance index (C-index) (32) for model ranking. Advanced users can supply custom-built preprocessors, models, and evaluation metrics. Automated diagnostics are integrated to identify common constraints in survival modeling, including low events-per-variable (EPV), outcome-associated missingness, and Cox-family assumption violations for proportional hazards (33) or log-hazard linearity (34). `mlsurv` also generates tiered model recommendations based on published best practice EPV and sample size thresholds (34–37).

Pre-processing follows a fixed stage order: imputation, scaling, and feature selection. The full preprocessing pipeline is fitted independently within each fold so that CV scores reflect out-of-sample performance of the complete pipeline, preventing the inflated estimates that arise when preprocessing uses leaked information from validation folds.

### Model Development

`mlsurv` provides two complementary approaches for building survival models. `run()` executes the full model development pipeline in a single call: training, tuning, test set evaluation, and bootstrap confidence intervals. Alternatively, the individual commands—`train()`, `tune()`, `evaluate()`, and `bootstrap()`—support incremental workflows that can be used to compare default and tuned models or add models to an existing comparison. By default, all models share identical preprocessing, CV folds, and metrics for fair head-to-head comparison.

Hyperparameter tuning uses Optuna’s Tree-structured Parzen Estimator (TPE) for Bayesian optimization (17) with default EPV-aware search space narrowing and convergence detection. The search space spans both model hyperparameters and preprocessing parameters, enabling joint optimization across the full model development pipeline. A single `random_state` parameter seeds all underlying frameworks (scikit-learn (14), XGBoost (13), PyTorch (38)) for end-to-end reproducibility.

### Model Evaluation

The `evaluate()` command measures the models’ performance on the held-out test set across discrimination and calibration metrics designed for right-censored data (**Table 3**). The scoring metric used for model selection is configured during the `setup()`command. The `bootstrap()` command derives confidence intervals by repeatedly measuring model performance on the test set through resampling (39). Pairwise model comparisons via the Kang nonparametric z-test (40) assess whether performance differences are statistically significant, and quantile-based risk stratification evaluates clinical utility via log-rank (41) and restricted mean survival time (RMST) (42,43) comparisons.

**Table 3.**
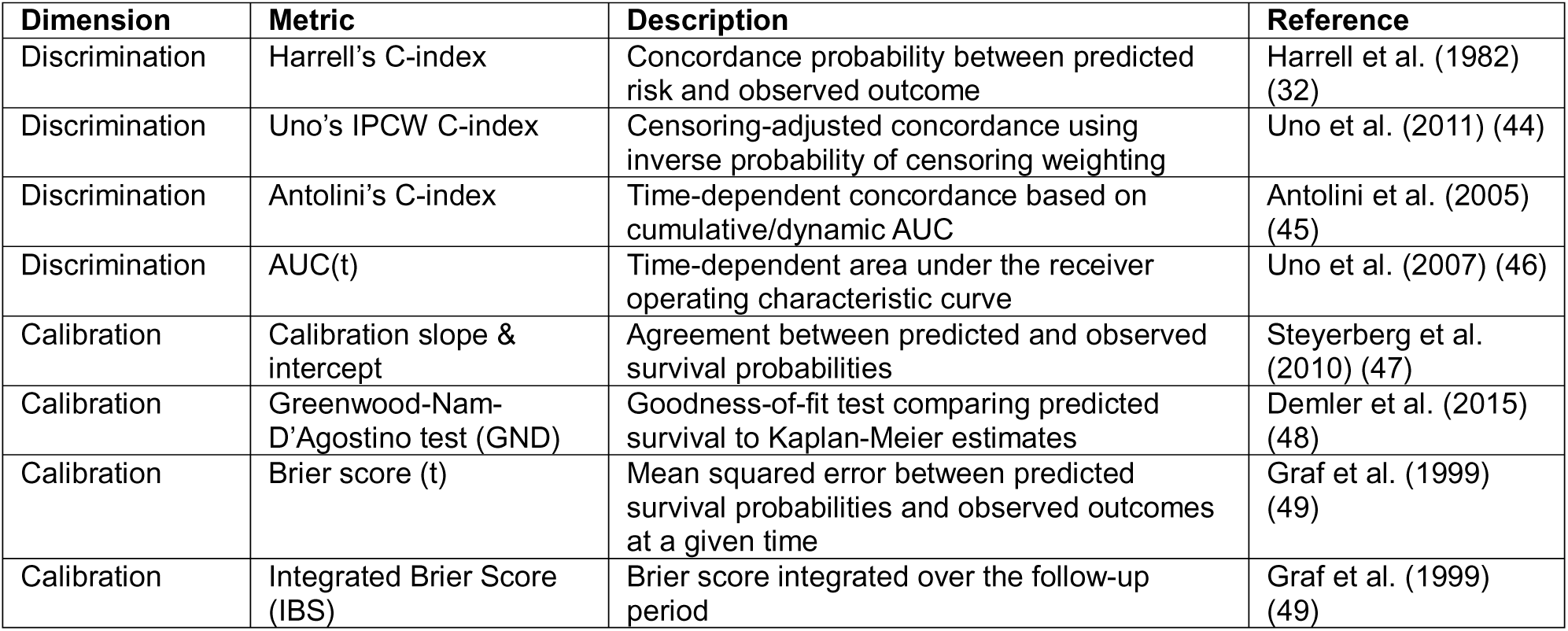
Evaluation metrics for right-censored survival data produced by mlsurv.

When cohort subpopulations are declared at `setup()`, `mlsurv` evaluates each model within every subpopulation across all metrics. Event by subpopulation stratification in splits and CV folds preserves subpopulation proportions to prevent imbalanced evaluation, and subpopulations with fewer than 30 events are flagged. When models show poor calibration, users are advised to test different available calibration strategies during model training.

For prospective use, the predict()command generates risk scores, survival probabilities at user-specified time points, and median survival times with bootstrap confidence intervals for new patient data beyond the existing dataset.

### Feature Analysis

Feature analysis quantifies which variables drive predictions and how they interact, spanning global importance across the cohort to local explanations for individual patients.

Three complementary methods quantify feature importance: permutation importance with significance testing via the permuted response method (50,51), applicable to all models; SHAP (Shapley Additive exPlanations) values, which decompose each prediction into per-feature contributions and aggregate to both global rankings and individual explanations (52); and Cox coefficients and hazard ratios with confidence intervals, for models that expose them.

Interaction analysis identifies pairs of variables whose joint influence on predicted risk differs from the sum of their separate contributions. The H-statistic quantifies interaction strength via partial dependence for all nonlinear models (53), and SHAP interaction values decompose pairwise contributions per-sample for tree-based models (52,54). Two-dimensional partial-dependence surfaces visualize the joint effect of top pairs.

At the individual level, `explain()` decomposes a prediction into per-feature SHAP contributions, rendered as an interactive waterfall plot. Explanations apply to new patient data as well as the evaluation set.

### Validation and Benchmarking

External validation on independent cohorts is essential for assessing whether a model generalizes beyond its training population (19,55). The `validate()`command evaluates all trained models on one or more held-out datasets, computing the same discrimination and calibration metrics used during model training.

Beyond external validation, `mlsurv` provides comparative benchmarking to quantify the added value of a new model over one or more established baselines such as clinical scores or single biomarkers. This is done by using the `benchmark()` method to compute delta C-index, Net Reclassification Improvement (NRI), and Integrated Discrimination Improvement (IDI) with bootstrap confidence intervals and significance testing (56,57).

### Comprehensive and Interactive Reporting

Throughout the analysis pipeline, `mlsurv` automatically evaluates fourteen limitation flags that surface potential methodological concerns (**S2 Table**). Each flag explains the limitation, why it matters clinically or statistically, and a concrete suggested action. The flags span sample adequacy, data quality, model diagnostics, and study design. A three-tier warning system controls visibility: methodological warnings always surface to the user, suggestion warnings appear once per issue, and internal warnings are suppressed by default but available for debugging.

The `generate_report()` command produces a comprehensive interactive HTML report assembling all analyses, with visualizations rendered as interactive Plotly figures and also available as standalone methods for custom analyses (**S3 Table**). For individual patient reporting, the `patient_report()` command produces an HTML with a predicted survival curve and summary, a SHAP feature explainer waterfall plot, and training set nearest-neighbor tables. Lastly, `export_results()` writes all results to spreadsheet, CSV, or Parquet formats, and `save_learner()` serializes the full learner state for later reuse.

The report includes a pre-populated TRIPOD+AI compliance checklist mapped against the 52- item reporting standard for AI-based prediction models (19). Of the 52 items, 18 are auto-filled from analysis metadata, 10 are scaffolded with guidance referencing the relevant report section, 22 are flagged as researcher-provided, and 2 are marked as not applicable. This simplifies the user’s reporting burden while ensuring that the computable items are populated accurately and consistently.

## Results and Discussion

### Three Design Principles for Biomedical Survival Machine Learning

We designed `mlsurv` around three core design principles that address key requirements in survival machine learning analysis. **Methodological rigor** ensures best practices for model development from pre-processing through hyperparameter tuning to protect against overfitting and data leakage. **Multi-scale evaluation and interpretation** enables comprehensive performance assessment of models across the full dataset, stratified subpopulations, and individual patients. Interpretation of the features used by a model or models is done using global importance and local explanations. **Automated trust and transparency** grounds `mlsurv`’s pipeline in established reporting standards through a tiered warning system for dataset and methodological limitations, structured outputs via an interactive report and comprehensive spreadsheet of results, and TRIPOD+AI reporting compliance. Seven tutorial vignettes on public datasets demonstrate one or more of these principles in use (**Table 4**). Vignette 7 demonstrates all three principles in an end-to-end analysis of the Chowell immunotherapy cohort (23) and its LORIS response scores (58).

**Table 4.**
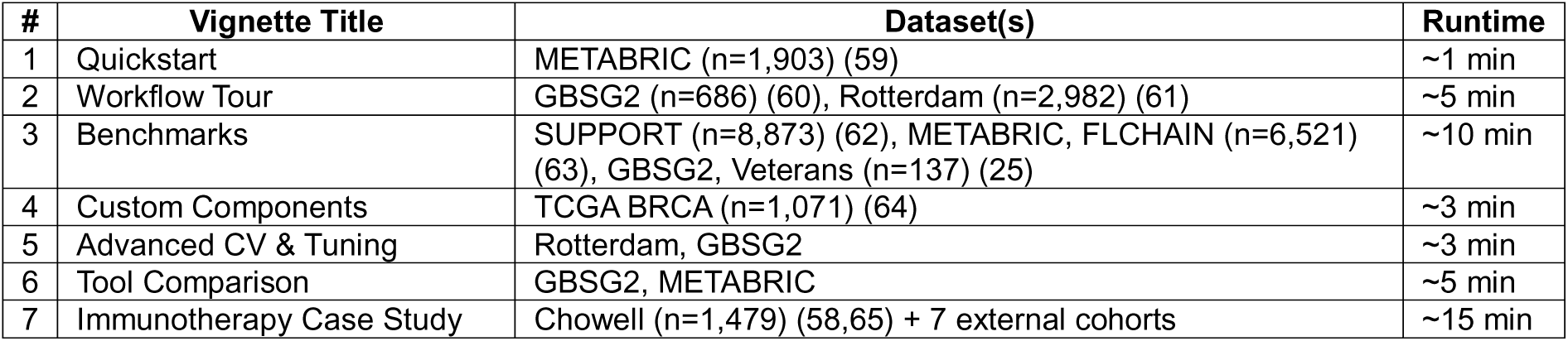
Vignettes showcasing mlsurv capabilities.

| # | Vignette Title | Dataset(s) | Runtime |
| --- | --- | --- | --- |
| 1 | Quickstart | METABRIC (n=1,903) (59) | ~1 min |
| 2 | Workflow Tour | GBSG2 (n=686) (60), Rotterdam (n=2,982) (61) | ~5 min |
| 3 | Benchmarks | SUPPORT (n=8,873) (62), METABRIC, FLCHAIN (n=6,521) (63), GBSG2, Veterans (n=137) (25) | ~10 min |
| 4 | Custom Components | TCGA BRCA (n=1,071) (64) | ~3 min |
| 5 | Advanced CV & Tuning | Rotterdam, GBSG2 | ~3 min |
| 6 | Tool Comparison | GBSG2, METABRIC | ~5 min |
| 7 | Immunotherapy Case Study | Chowell (n=1,479) (58,65) + 7 external cohorts | ~15 min |

#### Principle 1: Methodological Rigor

Survival ML models frequently overfit on their training cohort and fail to generalize to unseen cohorts. Overfitting can result from a mismatch between model complexity and dataset characteristics, information leakage across data splits, or inappropriate performance evaluation. `mlsurv` addresses these concerns with pre-training diagnostics, leakage-free pipelines, and censoring-aware discrimination and calibration metrics for model evaluation. With these safeguards, we used `mlsurv` to reproduce published benchmark C-indices with mean absolute deviation below 0.01 using default settings in Vignette 3 (**Table 5**).

**Table 5.** mlsurv performance against published concordance indices on five public datasets.

| Dataset | Model | `mlsurv` | Target | Gap | Source |
| --- | --- | --- | --- | --- | --- |
| GBSG2 | CoxNet | 0.668 | 0.669 | -0.001 | scikit-survival docs |
| GBSG2 | RSF | 0.690 | 0.691 | -0.001 | scikit-survival docs |
| Veterans | FSSVM | 0.724 | 0.720 | +0.004 | scikit-survival docs |
| METABRIC | CoxPH | 0.631 | 0.628* | +0.003 | Kvamme 2019 (66) |
| METABRIC | DeepSurv | 0.639 | 0.636* | +0.003 | Kvamme 2019 (66) |
| METABRIC | RSF | 0.662 | 0.649* | +0.013 | Kvamme 2019 (66) |
| METABRIC | DeepHit | 0.673 | 0.675* | -0.002 | Kvamme 2021 (15) |
| FLCHAIN | CoxPH | 0.794 | 0.790* | +0.004 | Kvamme 2019 (66) |
| FLCHAIN | DeepSurv | 0.796 | 0.790* | +0.006 | Kvamme 2019 (66) |
| FLCHAIN | RSF | 0.785 | 0.784* | +0.001 | Kvamme 2019 (66) |
| FLCHAIN | DeepHit | 0.794 | 0.791* | +0.003 | Kvamme 2021 (15) |
| SUPPORT | CoxPH | 0.595 | 0.598* | -0.003 | Kvamme 2019 (66) |
| SUPPORT | DeepSurv | 0.582 | 0.611* | -0.029 | Kvamme 2019 (66) |
| SUPPORT | DeepSurv | 0.611* (tuned) | 0.611* | +0.000 | Kvamme 2019 (66) |
| SUPPORT | RSF | 0.629 | 0.628* | +0.001 | Kvamme 2019 (66) |
| SUPPORT | DeepHit | 0.600 | 0.639* | -0.039 | Kvamme 2021 (15) |
| SUPPORT | DeepHit | 0.639* (tuned) | 0.639* | +0.000 | Kvamme 2021 (15) |

See Vignette 3 for full analysis. Gap = `mlsurv` performance – target, where a positive number indicates `mlsurv` outperformed the target. `mlsurv` models are computed using default hyperparameter settings unless marked with an asterisk (\*), indicating model hyperparameters were tuned. `mlsurv` tuned with `n_trials=100` via Optuna. Kvamme manuscripts used random search across 300 configurations for DeepSurv and DeepHit, grid search across 20 configurations for RSF, and L2 (ridge penalty) tuning across 9 values for CoxPH models. Mean absolute deviation (MAD, $\text{mean}(|\text{gap}|)$ ) was MAD=0.0076 using all `mlsurv` default settings and MAD=0.0031 when `mlsurv` tuned for deep learning models with SUPPORT.

#### Principle 2: Multi-Scale Evaluation and Interpretation

A key challenge when using ML survival models is understanding how the model is making predictions. Understanding which groups of patients benefit, which variables matter, and what drives an individual prediction are critical to developing new scientific hypotheses and using ML models well in clinical contexts. `mlsurv` evaluates models along two complementary axes: (1) the observation axis, from population to subpopulation to individual patients; and (2) the feature axis, with individual feature importances as well as feature interaction quantification. Vignette 2 presents a feature importance analysis along with patient-level explanations, and the Chowell immunotherapy case study in Vignette 7 discusses feature interaction and subpopulation analyses.

#### Principle 3: Automated Trust and Transparency

Methodological validation, transparent reporting, and reproducibility are essential but rarely built into ML survival software tools. `mlsurv` automates all three: fourteen limitation flags detect methodological concerns (**S2 Table**); a single `generate_report()` command assembles all results, limitations, environment details, and a pre-populated TRIPOD+AI reporting checklist in a standalone, interactive HTML report; lastly, `save_learner()` preserves the analysis state for full-state reproducibility. Vignettes 1, 2, 4, and 7 each produce a generated report. The Chowell case study in Vignette 7 demonstrates limitation flagging for non-proportional hazards and small subpopulation size warnings.

### Case Study: Immunotherapy Response Prediction

Immune checkpoint blockade (ICB) has emerged as a standard-of-care therapy across multiple cancer types (67–72), but as many as half of patients show little to no clinical benefit (73–80). Robust outcome prediction can help identify likely non-responders to avoid the toxicity, cost, and delay of ineffective therapy. We demonstrate the utility of `mlsurv` by predicting cancer patient outcomes when treated with ICB therapy through analysis of the Chowell immunotherapy cohort (23).

#### Cohort and Configuration

The Chowell immunotherapy cohort comprises 1,479 patients across 16 cancer types treated with ICB therapy at Memorial Sloan Kettering Cancer Center from 2015 to 2018 (23) (**Fig 2A**). Recently, a study used this cohort to develop the LORIS (logistic regression-based immunotherapy-response score) machine learning model to accurately predict patient response to ICB therapy using six variables: age, albumin, neutrophil-to-lymphocyte ratio (NLR), tumor mutational burden (TMB), systemic therapy history, and cancer type (58). Results from our survival models are compared to three references: tumor mutational burden (TMB) and PD-L1 expression, the two FDA-approved standard-of-care biomarkers used to predict patient response to ICB, and the published LORIS score for each patient. We train ML models to predict both overall survival (OS, **Fig 2**) and progression-free survival (PFS, **S1 Fig**) outcomes.

**Fig 2.**
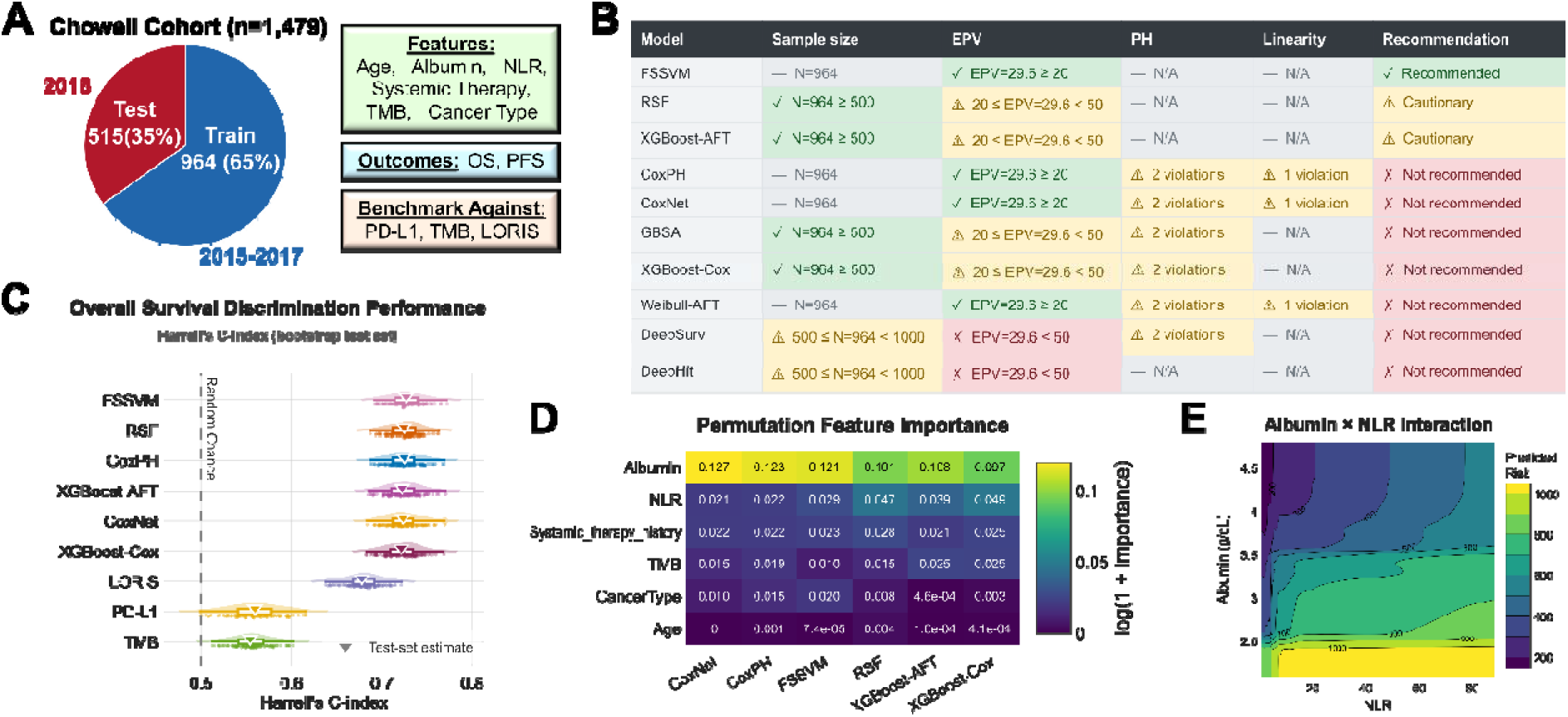
Chowell immunotherapy survival analysis. A) The Chowell immunotherapy cohort consists of 1,479 patients treated with ICB therapy. The dataset is temporally split into n=964 (65%) training observations from 2015 to 2017 and n=515 (35%) test observations from 2018 to match LORIS methods. Models are trained on age, albumin, neutrophil to lymphocyte ratio (NLR), systemic therapy history, tumor mutational burden (TMB), and cancer type. Two outcomes are assessed: overall survival (OS) and progression-free survival (PFS). Model performance is benchmarked against PD-L1 tumor proportion score, TMB, and the LORIS score. B) `setup()` runs a dataset audit for sample size, events per variable (EPV), and the Cox assumptions of proportional hazards (PH) and log-hazard linearity. The audit feeds into a model recommendations table which scores each `mlsurv` model as recommended, cautionary, or not recommended. Due to PH and linearity violations within the overall survival (OS) dataset as well as low EPV, only FSSVM is fully recommended, while RSF and XGBoost-AFT are noted as cautionary. C) `evaluate(bootstrap=True)` evaluates the discriminative performance of each OS model using Harrell’s concordance index. Bootstrapping the test set produces a performance distribution for each model and inverted triangles denote the performance of the full test set. All OS models show better performance versus benchmarks LORIS, PD-L1, and TMB. D) Permutation feature importance shows albumin consistently ranks as the most important feature for OS prediction followed by NLR. E) The partial dependence plot between albumin and NLR for the RSF model shows how predicted risk varies with the joint effects of both features. The highest risk exists where albumin drops below 2.5 g/dL and NLR is above 10.

The first step in our `mlsurv` pipeline is the `setup()`command. We use the LORIS temporal split—964 patients treated 2015-2017 for training, 515 treated in 2018 for testing—rather than `mlsurv`’s internal splitter to ensure consistency with the LORIS benchmark. `mlsurv` default preprocessing (SimpleImputer, StandardScaler, no feature selection) was used with 5-fold cross validation. Cancer type is registered at set up time as a subpopulation indicator, jointly stratifying the CV folds alongside event status and enabling downstream per-cancer-type evaluation.

Beyond configuring the preprocessing pipeline and CV strategy, `mlsurv`’s `setup()` command computes dataset summary statistics and runs methodological diagnostics to provide model recommendations (**Fig 2B, Fig S1**). Despite sharing the same features, splits, and pipeline configuration, the OS and PFS analyses flagged distinct methodological concerns driven by their different event counts (29.6 vs. 38.7 EPV, respectively) and time-to-event distributions. The OS analysis flagged proportional hazard (PH) violations on albumin and systemic therapy history plus a log-hazard linearity violation on NLR, while the PFS analysis flagged albumin and age for PH violations and showed no linearity concerns. Model recommendations shifted accordingly: on OS, CoxPH and CoxNet were not recommended due to combined PH and linearity violations, while on PFS the lack of linearity violations cleared these models to a cautionary status instead.

#### Model Development

Six models (CoxPH, CoxNet, RSF, XGBoost-Cox, XGBoost-AFT, and FSSVM) were fit with a single `learner.run()` command per outcome (OS, PFS). This command executes the full per-model `tune()`, `train()`, `evaluate()`, and `bootstrap()` workflow using the shared `setup()` configuration. Optimal hyperparameters are selected by the highest average C-index score achieved in 5-fold cross validation, and then final models are trained on the full training set. Each fitted model is then evaluated on the held-out test set with bootstrap confidence intervals across both discrimination and calibration metrics.

For models predicting OS, all six `mlsurv` models significantly exceed LORIS performance by an increase in Harrell’s C-index of ΔC=0.039–0.050 (bootstrap p≈0.02* for every model). Fast Survival Support Vector Machine (FSSVM (29)), the top-performing model, achieves C=0.728 versus LORIS’s C=0.678, while TMB and PD-L1 alone reach only C=0.554 and C=0.560 respectively (**Fig 2C**). Notably, FSSVM was the top recommended model at `setup()` due to Cox violations within the dataset. Its performance here demonstrates the value of matching dataset diagnostics to model assumptions and sample-size requirements.

All OS prediction models were more accurate than the corresponding PFS prediction models. The best PFS model CoxNet achieved C=0.674, with ΔC=-0.054 from the best OS model. A similar gap in performance has been reported in a related study, where OS prediction was more accurate than predicting binary clinical benefit defined by tumor response (81). These results highlight the distinction between prognostication of a patient’s overall survival versus prediction of progression during a treatment. In this case study and related work, prediction of progression during ICB therapy is a more challenging target than predicting OS.

PFS-trained `mlsurv` models show parity with LORIS. Four of six `mlsurv` models outperform LORIS though the differences are not statistically significant (best model CoxNet C=0.674 vs. LORIS C=0.664, Δc=0.009; bootstrap p=0.34). All `mlsurv` models and LORIS perform significantly better than TMB (C=0.576) and PD-L1 (C=0.564).

Subpopulation evaluation across all 16 cancer types reveals substantial model accuracy differences, in part due to small sample sizes. On OS, performance ranges from C≈0.90-0.98 (Breast, n=13) down to C≈0.55-0.62 (Hepatobiliary, n=16), while NSCLC, the largest subtype (n=207), achieves C≈0.69-0.71. Bootstrap confidence intervals reflect the uncertainty around performance estimates across each subtype and metric.

#### Feature Analysis and Interpretation

For the models predicting OS, albumin is the most important feature with a permutation importance of around 0.10-0.13 across all models, roughly 5x the next feature (**Fig 2D**). In models predicting PFS, feature analysis showed both TMB and albumin co-led with permutation importance of 0.04-0.06 each. This suggests that low albumin, a marker of cachexia and systemic inflammation, is more prognostic of overall survival, while TMB is more specifically associated with ICB response.

Feature interactions are also relatively consistent across models. Albumin × NLR is the top pair on OS in every model, showing that low albumin paired with high NLR predicts worse outcomes than their marginal effects would indicate (**Fig 2E**). High NLR may reflect tumor-driven neutrophilia, an immune dysregulation that impairs antitumor response.

#### External Validation

The `validate()` call evaluates every fitted model across one or more external cohorts. For our analysis, we validate against seven independent ICB cohorts and an OS-only negative control of non-ICB patients assembled in the LORIS study (58). Most cohorts show a modest discrimination drop from the internal test set Δc ≈ 0.04–0.10 across all models, consistent with expected cross-cohort variation.

On the negative control cohort of non-ICB patients (OS only), `mlsurv` models largely retain their accuracy. `mlsurv` models achieve a C-index range of 0.67-0.70, only 0.02-0.06 below ICB cohort performance, indicating that much of the discriminative ability reflects general prognostic signal rather than ICB-specific effects.

#### Reporting

The `generate_report()`command bundles all results into a standalone HTML. It includes a pre-populated TRIPOD+AI checklist (19) that provides full or partial information on 29 of 52 items, simplifying methodological reporting. Next, `patient_report()` produces a patient-specific report HTML. As an example, patient 208 from the hold-out test set is placed at the 91^st^ risk percentile of the training cohort (high risk) and shows a median predicted survival of 1.2 months [bootstrap 95% CI 1.0-1.9 months]. A SHAP waterfall plot shows individual feature contributions to the patient’s risk score and k-nearest neighbor tables by risk score and feature-space distance show similar patients within the training set.

#### Case Study Scientific Findings

Our case study is the first to compare OS and PFS survival ML models with a classification ML model for ICB outcome prediction in the large, public Chowell dataset. Using `mlsurv`, we find that OS prediction is more accurate than PFS prediction, in accordance with prior studies (81,82). OS predictions may be more accurate than PFS predictions for several reasons.

Progression is often challenging to measure well, leading to inexact measures that make prediction difficult. In addition, the Chowell dataset’s patient features may be better suited to prognostic (OS) prediction than therapy response prediction even though they were first used to predict response. Next, we showed that LORIS matches PFS model performance but is less accurate for OS prediction versus time-to-event OS models. These results suggest that ML classification models that predict therapy response cannot be used for predicting other aspects of patient outcomes. We also observed feature interaction effects between albumin and NLR in our OS prediction models, where the adverse prognostic effect of low albumin was amplified in patients with high NLR. Patients exhibiting both risk factors had substantially poorer OS than would be anticipated based on the independent effects of each biomarker, indicating a clinically meaningful interaction. Follow up work to externally validate this interaction and, if validated, understand its biological mechanisms would be valuable. Each of these findings were easily uncovered by simple `mlsurv` commands and automated comprehensive reporting.

### Future Development

Planned near-term development includes integration with the Galaxy computational workbench to broaden access to researchers with limited informatics skills and support reproducible workflow sharing (83–85). We also plan to expand methodological scope to support left/interval censoring and offer a wider set of recalibration methods. Long-term development will focus on addressing the limitations discussed in the previous paragraph. Support for multi-modal input expansion for imaging and free text will be developed using a combination of external embedding pipelines and native architectures. Federated and multi-site learning will be implemented to enable site-local training with aggregated coefficient updates, preserving data privacy while extending the effective training cohort across institutions.

### Limitations

`mlsurv` has three important limitations. First, `mlsurv` is intended primarily as a research tool. Translating a model developed in `mlsurv` into clinical use involves downstream steps that fall outside the scope of the software and are best addressed within institutional and regulatory processes. These steps include regulatory-grade model validation, model integration into clinical workflows with appropriate display of model predictions to guide clinical decisions, and post-deployment performance monitoring. Second, input data to `mlsurv` is currently limited to tabular format. Raw imaging and free-text data require external embedding pipelines to be converted into tabular features before entering the `mlsurv` workflow. Third, `mlsurv` assumes a single training cohort; federated and multi-site learning protocols are not supported within the current framework.

## Conclusions

Survival analysis is a foundational tool of biomedical research, and machine learning methods for survival analysis are increasingly used to capture high-dimensional and nonlinear patterns that classical Cox regression cannot. Conducting these analyses requires methodological rigor, multi-scale evaluation, and transparent reporting. Several Python software packages provide survival model implementations, but these are limited to model training and prediction, leaving the many surrounding workflow steps undefined. `mlsurv` closes this gap by integrating dataset diagnostics, model development and evaluation, methodological flagging, comprehensive interpretation, and automated reporting into a single workflow. We validated `mlsurv` against published results for numerous public datasets and demonstrated a mean absolute discrepancy below 0.01 in concordance index metrics. With our case study, we then demonstrated a full `mlsurv` pipeline through dataset auditing, comprehensive evaluation across models and subpopulations, feature importance and interaction analysis, external validation, and both cohort- and patient-level automated reporting.

`mlsurv` fills three critical gaps in the ecosystem of software tools for survival analysis: (1) ensuring methodological rigor through strong guardrails; (2) enabling systematic model benchmarking; and (3) mitigating risks associated with AI-generated code. A main gap filled by `mlsurv` is enabling researchers to perform rigorous end-to-end survival ML analysis. At every step, `mlsurv` automates the use of best practices that would otherwise demand expertise across disparate tools and methods. This includes use of automated data validity checks, model assumption tests, and optimization of all survival models. A second gap that `mlsurv` fills is performance comparison across many survival models, including multiple types of ML models. This benchmarking is an important but often ignored step in performance optimization, as no single model has emerged as the consistent best choice for survival analysis (20–22). `mlsurv` also supports custom models (see vignette 4), enabling model developers to compare the performance of new models to the existing suite of `mlsurv` models. The third gap that `mlsurv` addresses is potential methodological and coding errors that arise from the use of “vibe coding” with large language models and artificial intelligence (AI) agents (9,10,86,87). Researchers can vibe code analysis scripts in seconds, but these scripts can include methodological errors and skip important tests. `mlsurv` uses built-in best practices and automated reporting to mitigate these exact risks. For these reasons, we believe there is substantial value of best practice data analysis software packages like `mlsurv` in an era of AI-generated code.

## Data Availability

All data used in this study are available online at https://github.com/goeckslab/mlsurv/tree/main/examples/data. This study did not produce new data.

## List of Abbreviations

AFT: accelerated failure time
AI: Artificial Intelligence
AUC: area under the curve (in this case, the receiver operating characteristic curve)
C-index: concordance index
CLI: command line interface
CoxPH: Cox proportional hazards
CSV: comma-separated values
CV: cross-validation
EPV: events per variable
FLCHAIN: Free Light Chain dataset
FSSVM: Fast Survival Support Vector Machine
GBSA: Gradient Boosted Survival Analysis
GBSG2: German Breast Cancer Study Group 2 dataset
GND: Greenwood-Nam-D’Agostino test
HTML: hypertext markup language
IBS: integrated Brier score
ICB: immune checkpoint blockade
IDI: Integrated Discrimination Improvement
LORIS: logistic regression-based immunotherapy-response score
MAD: mean absolute deviation
METABRIC: Molecular Taxonomy of Breast Cancer International Consortium dataset
ML: machine learning
NLR: neutrophil-to-lymphocyte ratio
NRI: Net Reclassification Improvement
OS: overall survival
PD-L1: programmed death-ligand 1
PFS: progression-free survival
PH: proportional hazards
RMST: restricted mean survival time
RSF: Random Survival Forest
SHAP: Shapley Additive exPlanations
SUPPORT: Study to Understand Prognoses Preferences Outcomes and Risks of Treatment dataset
TCGA BRCA: The Cancer Genome Atlas Breast Invasive Carcinoma dataset
TMB: tumor mutational burden
TPE: Tree-structured Parzen Estimator
TRIPOD+AI: Transparent Reporting of a multivariable prediction model for Individual Prognosis Or Diagnosis, plus Artificial Intelligence

## Declarations

### Ethics approval and Consent to Participate

Not applicable.

### Consent for Publication

Not applicable.

### Availability of Data and Materials

All data underlying the findings in this study are publicly available and require no special access. This study used only previously published datasets; no new data were generated. All `mlsurv` source code is available at https://github.com/goeckslab/mlsurv. We provide cached data files for reproducibility of our analyses within our GitHub project repository at https://github.com/goeckslab/mlsurv/tree/main/examples/data. Dataset citations are documented in **Table 4** and within each vignette where they are used.

- Project name: `mlsurv`
- Project home page: https://github.com/goeckslab/mlsurv
- Operating system(s): Platform independent
- Programming language: Python, CLI
- Other requirements: Deep models are GPU-optional
- License: MIT
- Any restrictions to use by non-academics: None

### Competing Interests

The authors declare that they have no competing interests.

### Funding

This work was supported by the National Institutes of Health awards U24CA284167 and U24HG006620 (JG, www.nih.gov) as well as Moffitt Cancer Center (JG, www.moffitt.org).

### Author Contributions

AFP and JG conceived and designed the study. AFP, JQ, PCML, and KD created the software. AFP, INB, and TJ analyzed the data. AFP drafted the manuscript with edits from PCML and JG. All authors read and approved the final manuscript.

## Notes

### Competing Interest Statement

The authors have declared no competing interest.

### Author Declarations

Source data were openly available to the public before the initiation of the study. Cached copies are available on the project GitHub page (https://github.com/goeckslab/mlsurv) and citations are provided in Table 4 and within each vignette as applicable.

## References

1. Cox DR. Regression Models and Life-Tables. J R Stat Soc Ser B Methodol. 1972 Jan 1;34(2):187–202. doi:10.1111/j.2517-6161.1972.tb00899.x

2. Wiegrebe S, Kopper P, Sonabend R, Bischl B, Bender A. Deep learning for survival analysis: a review. Artif Intell Rev. 2024 Feb 19;57(3):65. doi:10.1007/s10462-023-10681-3

3. Abbasi AF, Asim MN, Ahmed S, Vollmer S, Dengel A. Survival prediction landscape: an in-depth systematic literature review on activities, methods, tools, diseases, and databases. Front Artif Intell. 2024;7:1428501. doi:10.3389/frai.2024.1428501 PubMed PMID: 39021434; PubMed Central PMCID: PMC11252047.

4. Wang P, Li Y, Reddy CK. Machine Learning for Survival Analysis: A Survey. ACM Comput Surv CSUR. 2019 Feb 27;51(6):110:1–110:36. doi:10.1145/3214306

5. Sartori F, Codicè F, Caranzano I, Rollo C, Birolo G, Fariselli P, et al. A Comprehensive Review of Deep Learning Applications with Multi-Omics Data in Cancer Research. Genes. 2025 Jun;16(6):648. doi:10.3390/genes16060648

6. Sidorova J, Lozano JJ. Review: Deep Learning-Based Survival Analysis of Omics and Clinicopathological Data. Inventions. 2024 Jun;9(3):59. doi:10.3390/inventions9030059

7. Waqas A, Tripathi A, Stewart P, Naeini M, Schabath MB, Rasool G. Embedding-based Multimodal Learning on Pan-Squamous Cell Carcinomas for Improved Survival Outcomes [Internet]. arXiv; 2024 [cited 2026 Jun 11]. Available from: http://arxiv.org/abs/2406.08521 doi:10.48550/arXiv.2406.08521

8. Cabitza F, Jurman G, Molinari F, Bellazzi R. Why almost all ML models for medicine are wrong-and what we need for evidence-based medical AI. Int J Med Inf. 2026 Oct 1;219:106538. doi:10.1016/j.ijmedinf.2026.106538

9. Messeri L, Crockett MJ. The uncritical adoption of AI in science is alarming — we urgently need guard rails. Nature. 2026 May;653(8115):675–6. doi:10.1038/d41586-026-01557-x

10. Ge Y, Mei L, Duan Z, Li T, Zheng Y, Wang Y, et al. A Survey of Vibe Coding with Large Language Models [Internet]. arXiv; 2025 [cited 2026 Jun 17]. Available from: http://arxiv.org/abs/2510.12399 doi:10.48550/arXiv.2510.12399

11. Davidson-Pilon C. lifelines: survival analysis in Python. J Open Source Softw. 2019 Aug 4;4(40):1317. doi:10.21105/joss.01317

12. Pölsterl S. scikit-survival: A Library for Time-to-Event Analysis Built on Top of scikit-learn. J Mach Learn Res. 2020;21(212):1–6.

13. Chen T, Guestrin C. XGBoost: A Scalable Tree Boosting System. In: Proceedings of the 22nd ACM SIGKDD International Conference on Knowledge Discovery and Data Mining [Internet]. New York, NY, USA: Association for Computing Machinery; 2016 [cited 2026 Apr 6]. p. 785–94. (KDD ’16). Available from: https://dl.acm.org/doi/10.1145/2939672.2939785 doi:10.1145/2939672.2939785

14. Pedregosa F, Varoquaux G, Gramfort A, Michel V, Thirion B, Grisel O, et al. Scikit-learn: Machine Learning in Python. J Mach Learn Res. 2011;12(85):2825–30. doi:10.48550/arXiv.1201.0490

15. Kvamme H, Borgan Ø. Continuous and discrete-time survival prediction with neural networks. Lifetime Data Anal. 2021 Oct;27(4):710–36. doi:10.1007/s10985-021-09532-6 PubMed PMID: 34618267; PubMed Central PMCID: PMC8536596.

16. Katzman JL, Shaham U, Cloninger A, Bates J, Jiang T, Kluger Y. DeepSurv: personalized treatment recommender system using a Cox proportional hazards deep neural network. BMC Med Res Methodol. 2018 Feb 26;18:24. doi:10.1186/s12874-018-0482-1 PubMed PMID: 29482517; PubMed Central PMCID: PMC5828433.

17. Akiba T, Sano S, Yanase T, Ohta T, Koyama M. Optuna: A Next-generation Hyperparameter Optimization Framework. In: Proceedings of the 25th ACM SIGKDD International Conference on Knowledge Discovery & Data Mining [Internet]. New York, NY, USA: Association for Computing Machinery; 2019 [cited 2026 Apr 6]. p. 2623–31. (KDD ’19). Available from: https://dl.acm.org/doi/10.1145/3292500.3330701 doi:10.1145/3292500.3330701

18. Kapoor S, Narayanan A. Leakage and the reproducibility crisis in machine-learning-based science. Patterns. 2023 Sep 8;4(9):100804. doi:10.1016/j.patter.2023.100804 PubMed PMID: 37720327; PubMed Central PMCID: PMC10499856.

19. Collins GS, Moons KGM, Dhiman P, Riley RD, Beam AL, Van Calster B, et al. TRIPOD+AI statement: updated guidance for reporting clinical prediction models that use regression or machine learning methods. BMJ. 2024 Apr 16;385:e078378. doi:10.1136/bmj-2023-078378 PubMed PMID: 38626948; PubMed Central PMCID: PMC11019967.

20. Zhang Y, Wong G, Mann G, Muller S, Yang JYH. SurvBenchmark: comprehensive benchmarking study of survival analysis methods using both omics data and clinical data. GigaScience. 2022 Jul 30;11:giac071. doi:10.1093/gigascience/giac071 PubMed PMID: 35906887; PubMed Central PMCID: PMC9338425.

21. Birolo G, Rossi I, Sartori F, Rollo C, Fariselli P, Sanavia T. Beyond Cox models: Assessing the performance of machine-learning methods in non-proportional hazards and non-linear survival analysis. Comput Biol Med. 2025 Nov;198(Pt B):111176. doi:10.1016/j.compbiomed.2025.111176 PubMed PMID: 41108906.

22. Huang Y, Bazzazzadehgan S, Li J, Arabshomali A, Li M, Bhattacharya K, et al. Comparison of machine learning methods versus traditional Cox regression for survival prediction in cancer using real-world data: a systematic literature review and meta-analysis. BMC Med Res Methodol. 2025 Oct 28;25(1):243. doi:10.1186/s12874-025-02694-z

23. Chowell D, Yoo SK, Valero C, Pastore A, Krishna C, Lee M, et al. Improved prediction of immune checkpoint blockade efficacy across multiple cancer types. Nat Biotechnol. 2022 Apr;40(4):499–506. doi:10.1038/s41587-021-01070-8 PubMed PMID: 34725502; PubMed Central PMCID: PMC9363980.

24. Simon N, Friedman JH, Hastie T, Tibshirani R. Regularization Paths for Cox’s Proportional Hazards Model via Coordinate Descent. J Stat Softw. 2011 Mar 9;39:1–13. doi:10.18637/jss.v039.i05

25. Kalbfleisch JD, Prentice RL. The Statistical Analysis of Failure Time Data [Internet]. John Wiley & Sons, Inc.; 2002. (Wiley Series in Probability and Statistics). Available from: https://onlinelibrary.wiley.com/doi/book/10.1002/9781118032985 doi:10.1002/9781118032985

26. Ishwaran H, Kogalur UB, Blackstone EH, Lauer MS. Random survival forests. Ann Appl Stat. 2008 Sep;2(3):841–60. doi:10.1214/08-AOAS169

27. Hothorn T, Bühlmann P, Dudoit S, Molinaro A, Van Der Laan MJ. Survival ensembles. Biostatistics. 2006 Jul 1;7(3):355–73. doi:10.1093/biostatistics/kxj011

28. Barnwal A, Cho H, Hocking T. Survival Regression with Accelerated Failure Time Model in XGBoost. J Comput Graph Stat. 2022 Oct 2;31(4):1292–302. doi:10.1080/10618600.2022.2067548

29. Pölsterl S, Navab N, Katouzian A. Fast Training of Support Vector Machines for Survival Analysis. In: Appice A, Rodrigues PP, Santos Costa V, Gama J, Jorge A, Soares C, editors. Machine Learning and Knowledge Discovery in Databases. Cham: Springer International Publishing; 2015. p. 243–59. doi:10.1007/978-3-319-23525-7_15

30. Katzman JL, Shaham U, Cloninger A, Bates J, Jiang T, Kluger Y. DeepSurv: personalized treatment recommender system using a Cox proportional hazards deep neural network. BMC Med Res Methodol. 2018 Feb 26;18(1):24. doi:10.1186/s12874-018-0482-1

31. Lee C, Zame W, Yoon J, Schaar M van der. DeepHit: A Deep Learning Approach to Survival Analysis With Competing Risks. Proc AAAI Conf Artif Intell. 2018 Apr 26;32(1). doi:10.1609/aaai.v32i1.11842

32. Harrell FE, Califf RM, Pryor DB, Lee KL, Rosati RA. Evaluating the yield of medical tests. JAMA. 1982 May 14;247(18):2543–6. PubMed PMID: 7069920.

33. Grambsch PM, Therneau TM. Proportional hazards tests and diagnostics based on weighted residuals. Biometrika. 1994 Sep 1;81(3):515–26. doi:10.1093/biomet/81.3.515

34. Harrell FE. Regression Modeling Strategies: With Applications to Linear Models, Logistic and Ordinal Regression, and Survival Analysis [Internet]. Cham: Springer International Publishing; 2015 [cited 2026 Apr 6]. (Springer Series in Statistics). Available from: https://link.springer.com/10.1007/978-3-319-19425-7 doi:10.1007/978-3-319-19425-7

35. Riley RD, Snell KI, Ensor J, Burke DL, Harrell FE, Moons KG, et al. Minimum sample size for developing a multivariable prediction model: PART II - binary and time-to-event outcomes. Stat Med. 2019 Mar 30;38(7):1276–96. doi:10.1002/sim.7992 PubMed PMID: 30357870; PubMed Central PMCID: PMC6519266.

36. van der Ploeg T, Austin PC, Steyerberg EW. Modern modelling techniques are data hungry: a simulation study for predicting dichotomous endpoints. BMC Med Res Methodol. 2014 Dec 22;14:137. doi:10.1186/1471-2288-14-137 PubMed PMID: 25532820; PubMed Central PMCID: PMC4289553.

37. Ogundimu EO, Altman DG, Collins GS. Adequate sample size for developing prediction models is not simply related to events per variable. J Clin Epidemiol. 2016 Aug;76:175–82. doi:10.1016/j.jclinepi.2016.02.031 PubMed PMID: 26964707; PubMed Central PMCID: PMC5045274.

38. Paszke A, Gross S, Massa F, Lerer A, Bradbury J, Chanan G, et al. PyTorch: An Imperative Style, High-Performance Deep Learning Library. In: Advances in Neural Information Processing Systems [Internet]. Curran Associates, Inc.; 2019 [cited 2026 Apr 6]. Available from: https://proceedings.neurips.cc/paper/2019/hash/bdbca288fee7f92f2bfa9f7012727740-Abstract.html doi:10.48550/arXiv.1912.01703

39. Efron B, Tibshirani RJ. An Introduction to the Bootstrap. New York: Chapman and Hall/CRC; 1994. 456 p. doi:10.1201/9780429246593

40. Kang L, Chen W, Petrick NA, Gallas BD. Comparing two correlated C indices with right-censored survival outcome: a one-shot nonparametric approach. Stat Med. 2015 Feb 20;34(4):685–703. doi:10.1002/sim.6370 PubMed PMID: 25399736; PubMed Central PMCID: PMC4314453.

41. Mantel N. Evaluation of survival data and two new rank order statistics arising in its consideration. Cancer Chemother Rep. 1966 Mar;50(3):163–70. PubMed PMID: 5910392.

42. Royston P, Parmar MKB. Restricted mean survival time: an alternative to the hazard ratio for the design and analysis of randomized trials with a time-to-event outcome. BMC Med Res Methodol. 2013 Dec 7;13:152. doi:10.1186/1471-2288-13-152 PubMed PMID: 24314264; PubMed Central PMCID: PMC3922847.

43. Uno H, Claggett B, Tian L, Inoue E, Gallo P, Miyata T, et al. Moving beyond the hazard ratio in quantifying the between-group difference in survival analysis. J Clin Oncol Off J Am Soc Clin Oncol. 2014 Aug 1;32(22):2380–5. doi:10.1200/JCO.2014.55.2208 PubMed PMID: 24982461; PubMed Central PMCID: PMC4105489.

44. Uno H, Cai T, Pencina MJ, D’Agostino RB, Wei LJ. On the C-statistics for evaluating overall adequacy of risk prediction procedures with censored survival data. Stat Med. 2011 May 10;30(10):1105–17. doi:10.1002/sim.4154 PubMed PMID: 21484848; PubMed Central PMCID: PMC3079915.

45. Antolini L, Boracchi P, Biganzoli E. A time-dependent discrimination index for survival data. Stat Med. 2005 Dec 30;24(24):3927–44. doi:10.1002/sim.2427 PubMed PMID: 16320281.

46. Uno H, Cai T, Tian L, Wei LJ. Evaluating Prediction Rules for t-Year Survivors With Censored Regression Models. J Am Stat Assoc. 2007 Jun 1;102(478):527–37. doi:10.1198/016214507000000149

47. Steyerberg EW, Vickers AJ, Cook NR, Gerds T, Gonen M, Obuchowski N, et al. Assessing the performance of prediction models: a framework for traditional and novel measures. Epidemiology. 2010 Jan;21(1):128–38. doi:10.1097/EDE.0b013e3181c30fb2 PubMed PMID: 20010215; PubMed Central PMCID: PMC3575184.

48. Demler OV, Paynter NP, Cook NR. Tests of calibration and goodness-of-fit in the survival setting. Stat Med. 2015 May 10;34(10):1659–80. doi:10.1002/sim.6428 PubMed PMID: 25684707; PubMed Central PMCID: PMC4555993.

49. Graf E, Schmoor C, Sauerbrei W, Schumacher M. Assessment and comparison of prognostic classification schemes for survival data. Stat Med. 1999 Sep 15;18(17– 18):2529–45. doi:10.1002/(sici)1097-0258(19990915/30)18:17/18<2529::aid sim274>3.0.co;2-5 PubMed PMID: 10474158.

50. Altmann A, Toloşi L, Sander O, Lengauer T. Permutation importance: a corrected feature importance measure. Bioinformatics. 2010 May 15;26(10):1340–7. doi:10.1093/bioinformatics/btq134 PubMed PMID: 20385727.

51. Breiman L. Random Forests. Mach Learn. 2001 Oct 1;45(1):5–32. doi:10.1023/A:1010933404324

52. Lundberg SM, Lee SI. A Unified Approach to Interpreting Model Predictions. In: Advances in Neural Information Processing Systems [Internet]. Curran Associates, Inc.; 2017 [cited 2026 Apr 6]. Available from: https://proceedings.neurips.cc/paper/2017/hash/8a20a8621978632d76c43dfd28b67767-Abstract.html doi:10.48550/arXiv.1705.07874

53. Friedman JH, Popescu BE. Predictive learning via rule ensembles. Ann Appl Stat. 2008 Sep;2(3):916–54. doi:10.1214/07-AOAS148

54. Lundberg SM, Erion G, Chen H, DeGrave A, Prutkin JM, Nair B, et al. From Local Explanations to Global Understanding with Explainable AI for Trees. Nat Mach Intell. 2020 Jan;2(1):56–67. doi:10.1038/s42256-019-0138-9 PubMed PMID: 32607472; PubMed Central PMCID: PMC7326367.

55. Steyerberg EW, Harrell FE. Prediction models need appropriate internal, internal-external, and external validation. J Clin Epidemiol. 2016 Jan;69:245–7. doi:10.1016/j.jclinepi.2015.04.005 PubMed PMID: 25981519; PubMed Central PMCID: PMC5578404.

56. Pencina MJ, D’Agostino RB, D’Agostino RB, Vasan RS. Evaluating the added predictive ability of a new marker: from area under the ROC curve to reclassification and beyond. Stat Med. 2008 Jan 30;27(2):157–72; discussion 207-212. doi:10.1002/sim.2929 PubMed PMID: 17569110.

57. Pencina MJ, D’Agostino RB, Steyerberg EW. Extensions of net reclassification improvement calculations to measure usefulness of new biomarkers. Stat Med. 2011 Jan 15;30(1):11–21. doi:10.1002/sim.4085 PubMed PMID: 21204120; PubMed Central PMCID: PMC3341973.

58. Chang TG, Cao Y, Sfreddo HJ, Dhruba SR, Lee SH, Valero C, et al. LORIS robustly predicts patient outcomes with immune checkpoint blockade therapy using common clinical, pathologic and genomic features. Nat Cancer. 2024 Aug;5(8):1158–75. doi:10.1038/s43018-024-00772-7

59. Curtis C, Shah SP, Chin SF, Turashvili G, Rueda OM, Dunning MJ, et al. The genomic and transcriptomic architecture of 2,000 breast tumours reveals novel subgroups. Nature. 2012 Jun;486(7403):346–52. doi:10.1038/nature10983

60. Schumacher M, Bastert G, Bojar H, Hübner K, Olschewski M, Sauerbrei W, et al. Randomized 2 x 2 trial evaluating hormonal treatment and the duration of chemotherapy in node-positive breast cancer patients. German Breast Cancer Study Group. J Clin Oncol Off J Am Soc Clin Oncol. 1994 Oct;12(10):2086–93. doi:10.1200/JCO.1994.12.10.2086 PubMed PMID: 7931478.

61. Royston P, Altman DG. External validation of a Cox prognostic model: principles and methods. BMC Med Res Methodol. 2013 Mar 6;13(1):33. doi:10.1186/1471-2288-13-33

62. Knaus WA, Harrell FE, Lynn J, Goldman L, Phillips RS, Connors AF, et al. The SUPPORT prognostic model. Objective estimates of survival for seriously ill hospitalized adults. Study to understand prognoses and preferences for outcomes and risks of treatments. Ann Intern Med. 1995 Feb 1;122(3):191–203. doi:10.7326/0003-4819-122-3-199502010-00007 PubMed PMID: 7810938.

63. Dispenzieri A, Katzmann JA, Kyle RA, Larson DR, Therneau TM, Colby CL, et al. Use of nonclonal serum immunoglobulin free light chains to predict overall survival in the general population. Mayo Clin Proc. 2012 Jun;87(6):517–23. doi:10.1016/j.mayocp.2012.03.009 PubMed PMID: 22677072; PubMed Central PMCID: PMC3538473.

64. Koboldt DC, Fulton RS, McLellan MD, Schmidt H, Kalicki-Veizer J, McMichael JF, et al. Comprehensive molecular portraits of human breast tumours. Nature. 2012 Oct;490(7418):61–70. doi:10.1038/nature11412

65. Chowell D, Morris LGT, Grigg CM, Weber JK, Samstein RM, Makarov V, et al. Patient HLA class I genotype influences cancer response to checkpoint blockade immunotherapy. Science. 2018 Feb 2;359(6375):582–7. doi:10.1126/science.aao4572

66. Kvamme H, Borgan Ø, Scheel I. Time-to-Event Prediction with Neural Networks and Cox Regression. J Mach Learn Res. 2019;20(129):1–30. doi:10.48550/arXiv.1907.00825

67. Sharma P, Goswami S, Raychaudhuri D, Siddiqui BA, Singh P, Nagarajan A, et al. Immune checkpoint therapy-current perspectives and future directions. Cell. 2023 Apr 13;186(8):1652–69. doi:10.1016/j.cell.2023.03.006 PubMed PMID: 37059068.

68. Goleva E, Lyubchenko T, Kraehenbuehl L, LaCouture ME, Leung DYM, Kern JA. Our Current Understanding of Checkpoint Inhibitor Therapy in Cancer Immunotherapy. Ann Allergy Asthma Immunol Off Publ Am Coll Allergy Asthma Immunol. 2021 Jun;126(6):630–8. doi:10.1016/j.anai.2021.03.003 PubMed PMID: 33716146; PubMed Central PMCID: PMC8713301.

69. Reck M, Rodríguez-Abreu D, Robinson AG, Hui R, Csőszi T, Fülöp A, et al. Five-Year Outcomes With Pembrolizumab Versus Chemotherapy for Metastatic Non–Small-Cell Lung Cancer With PD-L1 Tumor Proportion Score ≥ 50%. J Clin Oncol. 2021 Jul 20;39(21):2339–49. doi:10.1200/JCO.21.00174

70. Brahmer JR, Tykodi SS, Chow LQM, Hwu WJ, Topalian SL, Hwu P, et al. Safety and Activity of Anti–PD-L1 Antibody in Patients with Advanced Cancer. N Engl J Med. 2012 Jun 28;366(26):2455–65. doi:10.1056/NEJMoa1200694

71. Immune Checkpoint Blockade plus Axitinib for Renal-Cell Carcinoma. N Engl J Med. 2019 Jun 27;380(26):2581–2. doi:10.1056/NEJMc1905518

72. Cercek A, Lumish M, Sinopoli J, Weiss J, Shia J, Lamendola-Essel M, et al. PD-1 Blockade in Mismatch Repair–Deficient, Locally Advanced Rectal Cancer. N Engl J Med. 2022 Jun 22;386(25):2363–76. doi:10.1056/NEJMoa2201445

73. Postow MA, Sidlow R, Hellmann MD. Immune-Related Adverse Events Associated with Immune Checkpoint Blockade. N Engl J Med. 2018 Jan 11;378(2):158–68. doi:10.1056/NEJMra1703481

74. Haslam A, Olivier T, Prasad V. How many people in the US are eligible for and respond to checkpoint inhibitors: An empirical analysis. Int J Cancer. 2025 Jun 15;156(12):2352–9. doi:10.1002/ijc.35347 PubMed PMID: 39887747.

75. Haslam A, Prasad V. Estimation of the Percentage of US Patients With Cancer Who Are Eligible for and Respond to Checkpoint Inhibitor Immunotherapy Drugs. JAMA Netw Open. 2019 May 3;2(5):e192535. doi:10.1001/jamanetworkopen.2019.2535 PubMed PMID: 31050774; PubMed Central PMCID: PMC6503493.

76. Pilard C, Ancion M, Delvenne P, Jerusalem G, Hubert P, Herfs M. Cancer immunotherapy: it’s time to better predict patients’ response. Br J Cancer. 2021 Sep;125(7):927–38. doi:10.1038/s41416-021-01413-x PubMed PMID: 34112949; PubMed Central PMCID: PMC8476530.

77. Havel JJ, Chowell D, Chan TA. The evolving landscape of biomarkers for checkpoint inhibitor immunotherapy. Nat Rev Cancer. 2019 Mar;19(3):133–50. doi:10.1038/s41568-019-0116-x PubMed PMID: 30755690; PubMed Central PMCID: PMC6705396.

78. Wolchok JD, Chiarion-Sileni V, Rutkowski P, Cowey CL, Schadendorf D, Wagstaff J, et al. Final, 10-Year Outcomes with Nivolumab plus Ipilimumab in Advanced Melanoma. N Engl J Med. 2025 Jan 1;392(1):11–22. doi:10.1056/NEJMoa2407417

79. Gandhi L, Rodríguez-Abreu D, Gadgeel S, Esteban E, Felip E, Angelis FD, et al. Pembrolizumab plus Chemotherapy in Metastatic Non–Small-Cell Lung Cancer. N Engl J Med. 2018 May 31;378(22):2078–92. doi:10.1056/NEJMoa1801005

80. Tannir NM, Albigès L, McDermott DF, Burotto M, Choueiri TK, Hammers HJ, et al. Nivolumab plus ipilimumab versus sunitinib for first-line treatment of advanced renal cell carcinoma: extended 8-year follow-up results of efficacy and safety from the phase III CheckMate 214 trial. Ann Oncol Off J Eur Soc Med Oncol. 2024 Nov;35(11):1026–38. doi:10.1016/j.annonc.2024.07.727 PubMed PMID: 39098455; PubMed Central PMCID: PMC11907766.

81. Yoo SK, Fitzgerald CW, Cho BA, Fitzgerald BG, Han C, Koh ES, et al. Prediction of checkpoint inhibitor immunotherapy efficacy for cancer using routine blood tests and clinical data. Nat Med. 2025 Mar;31(3):869–80. doi:10.1038/s41591-024-03398-5 PubMed PMID: 39762425; PubMed Central PMCID: PMC11922749.

82. Li Y, Brendel M, Wu N, Ge W, Zhang H, Rietschel P, et al. Machine learning models for identifying predictors of clinical outcomes with first-line immune checkpoint inhibitor therapy in advanced non-small cell lung cancer. Sci Rep. 2022 Oct 21;12(1):17670. doi:10.1038/s41598-022-20061-6

83. The Galaxy Community. Galaxy for accessible, reproducible, and collaborative data analyses: 2026 update. Nucleic Acids Res. 2026 Jun 9;gkag469. doi:10.1093/nar/gkag469

84. Giardine B, Riemer C, Hardison RC, Burhans R, Elnitski L, Shah P, et al. Galaxy: A platform for interactive large-scale genome analysis. Genome Res. 2005 Oct;15(10):1451–5. doi:10.1101/gr.4086505 PubMed PMID: 16169926; PubMed Central PMCID: PMC1240089.

85. Goecks J, Nekrutenko A, Taylor J, The Galaxy Team. Galaxy: a comprehensive approach for supporting accessible, reproducible, and transparent computational research in the life sciences. Genome Biol. 2010 Aug 25;11(8):R86. doi:10.1186/gb-2010-11-8-r86

86. Moore JH, Tatonetti N. Vibe coding: a new paradigm for biomedical software development. BioData Min. 2025 Jul 1;18(1):46. doi:10.1186/s13040-025-00462-9

87. Lee Y, Huh S. How Can Clinicians Leverage Vibe Coding for Machine Learning and Deep Learning Research? Endocrinol Metab. 2025 Oct;40(5):659–67. doi:10.3803/EnM.2025.2675 PubMed PMID: 41208262; PubMed Central PMCID: PMC12602019.

