## Supplemental Figure and Tables for "A software package for simple and rigorous survival machine learning analysis in biomedical research"

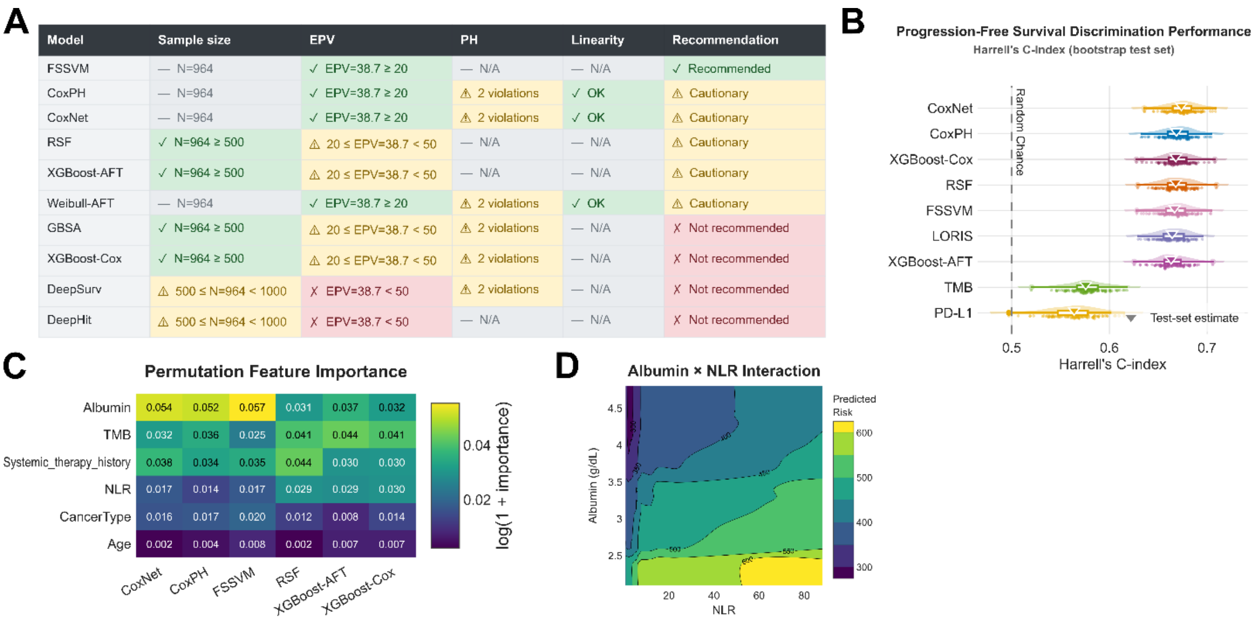

| Model | Sample size | EPV | PH | Linearity | Recommendation |
|---|---|---|---|---|---|
| FSSVM | — N=964 | ✓ EPV=38.7 ≥ 20 | — N/A | — N/A | ✓ Recommended |
| CoxPH | — N=964 | ✓ EPV=38.7 ≥ 20 | ⚠ 2 violations | ✓ OK | ⚠ Cautionary |
| CoxNet | — N=964 | ✓ EPV=38.7 ≥ 20 | ⚠ 2 violations | ✓ OK | ⚠ Cautionary |
| RSF | ✓ N=964 ≥ 500 | ⚠ 20 ≤ EPV=38.7 < 50 | — N/A | — N/A | ⚠ Cautionary |
| XGBoost-AFT | ✓ N=964 ≥ 500 | ⚠ 20 ≤ EPV=38.7 < 50 | — N/A | — N/A | ⚠ Cautionary |
| Weibull-AFT | — N=964 | ✓ EPV=38.7 ≥ 20 | ⚠ 2 violations | ✓ OK | ⚠ Cautionary |
| GBSA | ✓ N=964 ≥ 500 | ⚠ 20 ≤ EPV=38.7 < 50 | ⚠ 2 violations | — N/A | ✗ Not recommended |
| XGBoost-Cox | ✓ N=964 ≥ 500 | ⚠ 20 ≤ EPV=38.7 < 50 | ⚠ 2 violations | — N/A | ✗ Not recommended |
| DeepSurv | ⚠ 500 ≤ N=964 < 1000 | ✗ EPV=38.7 < 50 | ⚠ 2 violations | — N/A | ✗ Not recommended |
| DeepHit | ⚠ 500 ≤ N=964 < 1000 | ✗ EPV=38.7 < 50 | — N/A | — N/A | ✗ Not recommended |



**S1 Fig. Chowell immunotherapy survival analysis of progression-free survival**. A) `setup()` runs a dataset audit for sample size, events per variable (EPV), and the Cox assumptions of proportional hazards (PH) and log-hazard linearity. The audit feeds into a model recommendations table which scores each `mlsurv` model as recommended, cautionary, or not recommended. Due to PH violations and low EPV within this dataset, only FSSVM is fully recommended, while CoxPH, CoxNet, RSF, XGBoost-AFT, and Weibull-AFT are noted as cautionary. B) `evaluate(bootstrap=True)` evaluates the discriminative performance of each model using Harrell's concordance index. Bootstrapping the test set produces a performance distribution for each model and inverted triangles denote the performance of the full test set. All progression-free survival (PFS) models show similar performance to the LORIS score and improved performance versus benchmarks PD-L1 and TMB. C) Permutation feature importance shows albumin consistently ranks as the most important feature for PFS prediction followed by tumor mutational burden (TMB) and prior systemic therapy. These results contrast with the OS models which showed less importance of TMB and systemic therapy history and greater importance of albumin. D) The partial dependence plot between albumin and neutrophil to lymphocyte ratio (NLR) for the RSF model shows how predicted risk varies with the joint effects of both features. The highest risk exists where albumin drops below 2.5 g/dL and NLR is above 50.

**S1 Table. `mlsurv` high-level functions.**

| Method | Description |
|---|---|
| `setup()` | Configures pre-processing, data splitting, and random state. Audits dataset and recommends appropriate models. |
| `run()` | Runs the full train, tune, evaluate, bootstrap survival analysis pipeline. |
| `train()` | Trains one or more models with default or tuned hyperparameters. |
| `tune()` | Performs hyperparameter optimization using Optuna. |
| `evaluate()` | Evaluates trained models on the test set, optionally with AUC(t), feature importance, SHAP, and feature interactions. |
| `bootstrap()` | Computes bootstrap confidence intervals for evaluation metrics. |
| `summary()` | Returns a pandas data frame of all CV and test set evaluation metric results. |
| `validate()` | Evaluates an external validation cohort on one or more models. |
| `predict()` | Predicts survival for new patients from one or more models. |
| `explain()` | Explains an individual's risk prediction via SHAP values. |
| `benchmark()` | Compares model performance against reference scores. |
| `importance()` | Computes feature importance values, either permutation or SHAP. |
| `interactions()` | Computes pairwise feature interactions, H-statistic or SHAP. |
| `feature_summary()` | Outputs a table of combined feature importance values (permutation, SHAP, coef) |
| `generate_report()` | Generates a comprehensive analysis report. |
| `patient_report()` | Generates a single patient prediction report. |
| `ph_test()` | Runs the proportional hazards assumption test. |
| `linearity_test()` | Runs the log-hazard linearity test. |
| `rmst_test()` | Compares restricted mean survival time across risk groups. |
| `logrank_test()` | Compares survival curves across risk groups. |
| `importance_test()` | Computes feature importance p-values. |
| `parameter_sweep()` | Sweeps over model or preprocessing parameter ranges and evaluates each. |
| `list_studies()` | Lists all available Optuna studies. |
| `export_study()` | Export's a model's Optuna tuning history to CSV or JSON. |
| `reload_study()` | Reloads an Optuna study from disk. |
| `save_learner()` | Saves the entire learner state to a single file. |
| `load_learner()` | Loads a saved learner from disk. |
| `save_artifact()` | Saves a model artifact without associated training/test data, for privacy/size. |
| `load_artifact()` | Loads a saved artifact from disk. |
| `export_results()` | Exports all available learner results to file. |

### S2 Table. Automatic limitation flags evaluated by `mlsurv`.

| Flag | Trigger | Risk | Guidance |
|---|---|---|---|
| Small sample size | N < 200 (train + test) | Increases overfitting risk; unstable performance estimates | Collect more data, use simpler models, apply regularization |
| Low Events Per Variable (EPV) | EPV < 10 | Model parameters unreliably estimated; overfitting risk increases | Reduce features, use penalized models, or report results as exploratory |
| High censoring rate | Censoring > 80% | Reduces effective sample size for event-based metrics; may bias survival estimates toward longer follow-up | Verify follow-up accuracy; use IPCW-based metrics |
| Extensive missing data | Missing > 20% OR any variable > 50% | Bias if not missing completely at random; imputation assumptions may not hold | Investigate missingness mechanism; consider sensitivity analyses or excluding variables >50% missing |
| Proportional Hazards (PH) violations | Grambsch-Therneau FDR-adjusted p < 0.05 | Cox-family models assume constant hazard ratios; violations may bias predictions | Use non-PH models |
| Log-hazard linearity violations | RCS Wald FDR-adjusted p < 0.05 | Cox-family models assume linear effects on the log-hazard scale; non-linearity may bias hazard ratios and predictions | Apply non-linear transformations (e.g. restricted cubic splines, RCS) or switch to tree-ensemble or deep learning models |
| Train/test performance gap | CV mean - test C-index > 0.05 | Suggests overfitting | Increase regularization, reduce model complexity, or reduce tuning trials |
| Low discrimination | Test C-index < 0.6 | Near-random discrimination; features may lack sufficient prognostic information | Review features/data quality; try alternative models or predictors |
| Poor calibration | Slope outside [0.5, 1.5] | Systematic over-/under-prediction of risk | Investigate whether miscalibration is systematic; consider recalibration before deployment |
| Substantial multicollinearity | Any pair \|R\| > 0.9 OR 5 pairs \|R\| ≥ 0.8 | Inflates coefficient variance; distorts permutation importance; feature effects harder to disentangle | Remove or combine correlated features, use VIF selection |
| No external validation | No external cohort provided | Internal validation may overestimate performance | Validate on independent dataset; if unavailable, state this limitation in reporting |
| Small subpopulations | Any subpopulation with < 30 events | Performance estimates unreliable | Pool subpopulations or collect more data; report with caution |
| Model appropriateness | Trained model not recommended for dataset size/EPV | High overfitting risk for complex models | Switch to recommended models or collect more data |
| IPCW training-support violation | Test follow-up beyond censoring-distribution ($\hat{G}$) support | Inflated point estimates and bootstrap CIs for all IPCW-weighted metrics (late-time observations dominate) | Prefer Harrell/Antolini concordance for discrimination; pass `ipcw_tau` to override default truncation |

**S3 Table. Programmatic visualization methods available in `mlsurv`.**

| Method | Description |
|---|---|
| `plot_metric` | Model performance comparison with bootstrap CIs (forest, bar, raincloud, box, violin) |
| `plot_survival` | Kaplan-Meier curves, overall or by risk group with log-rank test and number-at-risk table |
| `plot_calibration` | Calibration curves (predicted vs observed survival) at a select time point with Greenwood formula confidence intervals |
| `plot_roc` | Time-dependent ROC curve at a select time point with bootstrap CI ribbons |
| `plot_metric_time` | Time-dependent AUC or Brier score curves |
| `plot_metric_heatmap` | Multi-metric heatmap across models |
| `plot_importance` | Permutation feature importance across models (bar, forest, raincloud, boxplot, heatmap) |
| `plot_shap` | SHAP values (importance bar, beeswarm) |
| `plot_interactions` | Feature interaction heatmap (H-statistic or SHAP) |
| `plot_interaction_pdp` | 2D partial dependence contour surface for feature interaction pairs |
| `plot_coefficients` | Cox coefficients (bar, forest) |
| `plot_coefficient_comparison` | Scatter plot comparing coefficients between models |
| `plot_hazard_ratios` | Hazard ratio forest (univariate, multivariate) |
| `plot_hazard_ratios_comparison` | Univariate vs multivariate hazard ratio comparison |
| `plot_diagnostics` | Diagnostic residuals (Schoenfeld for PH testing, martingale for log-hazard linearity assessment, deviance for outlier detection) |
| `plot_calibration_overlay` | Predicted vs. observed survival curves |
| `plot_benchmark_incremental` | Forest plot of delta-C, NRI, IDI for benchmark evaluation |
| `plot_feature_correlation` | Feature correlation heatmap |
| `plot_feature_distributions` | Feature distributions for train and test sets |
| `plot_missing` | Missing data heatmap |
| `plot_optuna` | Optuna optimization history and parameter importance |

Subpopulation-stratified and external-validation variants are available via view proxies: `learner.subpopulation(group).plot_*(...)` and `learner.validation(cohort).plot_*(...)`.
